# Covid-19 in Chile. The experience of a Regional reference Center. Preliminary report

**DOI:** 10.1101/2020.06.14.20130898

**Authors:** Felipe Olivares, Daniel Muñoz, Alberto Fica, Ignacio Delama, Ignacia Alvarez, Maritza Navarrete, Eileen Blackburn, Pamela Garrido, Juan Grandjean

**Author notes:** Correspondence to: Dr.Alberto Fica Cubillos, Médico Infectólogo, SubDepartamento de Medicina, Hospital Base de Valdivia, Bueras 1003, Valdivia, Región De Los Ríos, Chile, Instituto de Medicina, Facultad de Medicina, Campus Isla Teja, Universidad Austral de Chile, Valdivia, Chile, / Teléfono. Conflict of interest None / Funding: None. Título en español: Covid-19 en Chile. Experiencia en un centro de referencia regional. Reporte preliminar.

## Abstract

During the first pandemic wave Covid-19 reached Latin America cities.

**Aim:** To report clinical features and outcomes associated to Covid-19 in a group of patients admitted during the first wave in a regional reference Center in southern Chile designated to severe and critical cases.

**Methods:** Cases were identified by a compatible clinical picture associated to positive RT-PCR or serological testing. A standard protocol was applied.

**Results:** 21 adult patients (20 diagnosed by PCR, one by serology) were admitted between epidemiological weeks 13 to 20, involving 8.8% of total regional cases. Hospitalization occurred at a median of 11 days after symptoms onset. Patients ≥60 years old predominated (57.1%). Hypertension (61.9%), obesity (57.1%) and diabetes mellitus 2 (38.1%) were prevalent but 19% had no comorbid conditions nor were elderly. Two cases involved second-trimester pregnant women. Positive IgM or IgM/IgG results obtained by rapid serological testing were limited (19% at 1st week; 42.9% at 2nd week). Nine patients (42.9%, critical group) were transferred to ICU and connected to mechanical ventilation due to respiratory failure. By univariate analysis admission to ICU was significantly associated to tachypnea and higher plasmatic LDH values. One pregnant woman required urgent cesarean section given birth to a premature neonate without vertical transmission. Two patients died (in-hospital mortality 9.5%) and length of stay was ≥ 14 days in 57.9% of patients.

**Conclusion:** In our regional Center, Covid 19 was associated to known risk factors, had a prolonged stay and in-hospital mortality. Tachypnea ≥30/min is predictive of transfer to ICU.

## Introduction

Covid-19 is an emerging disease caused by the SARS-CoV-2 virus and first reported in late 2019 in China (1). Due to its rapid expansion, on March 2020 it was declared a global pandemic by the WHO (2) and up to date has affected more than 6 million habitants worldwide of whom 379,941 have died (3). The first case reported in Chile was registered on March 3^rd^ and at the beginning of June 105,000 cases has been reported with more than a thousand people deceased (4). The temporal evolution of this epidemic is unknown, but it has been estimated that it could generate recurrent outbreaks and remain in circulation for a long time (5). Currently there is little information on the clinical presentation of Covid-19 and outcomes in Latin America, especially in hospitalized patients (6-9). This could contrast with that reported in countries of the northern hemisphere, which have dissimilar features on demographic, economic, cultural, healthcare systems and/or mitigation strategies issues (10-19). We present this preliminary report on a cohort of Covid-19 patients during the first wave in a southern region in Chile detailing clinical features and outcomes in a reference center.

## Patients and Methods

### Study design, inclusion criteria and laboratory tests

Retrospective descriptive study of patients admitted during the first wave at the Hospital Regional of Valdivia, in southern Chile. Patients were included, if had suspicious symptoms plus either a positive SARS-CoV-2 RT-PCR or a rapid serological test coupled with compatible chest-CT imaging. For RT-PCR tests, nasopharyngeal swab samples were obtained and placed in viral transport medium. Total nucleic acids were extracted from 200 µL of transport medium with Chemagic Prepito Instrument using Prepito Viral DNA / RNA Kit (Both PerkinElmer ™) according to manufacturer’s recommendations and eluted in 100 µL of an elution buffer. SARS-CoV-2 RNA was detected using the Primerdesign COVID-19 Genesig Real-Time PCR assay and the amplification reaction was performed in a Bio-Rad® CFX96 thermocycler according to manufacturer’s instructions. Specimens were considered positive if the Ct value was <37 and the internal controls were detected. In parallel, the quality of the samples was checked with a human RNAse P Real-Time PCR assay with the CDC primers from IDT. Serology was performed using the “VivaDiag COVID □ 19 IgM / IgG Rapid Test lateral flow immunoassay (LFIA)” kit based on the addition of 10 µL of serum and then 2 to 3 drops of (70-100 µL) buffer dilution. Results were read after 15 minutes.

### Regional management protocol

Los Ríos Region has an estimated population of 406,000 inhabitants (20). Most of the population (> 80%) is cared for in an integrated network of primary care centers and low-complexity hospitals, with only one high-complexity reference center (Hospital Regional) designated to receive severe and critically ill Covid-19 patients. Severity illness stratification was performed based on a regional clinical care guideline using the following classification: a) Mild: no need of oxygen therapy or hospitalization, b) Moderate: low oxygen requirement (FiO2 <32%); PaO_2_ / FiO_2_ ratio ≥ 200, respiratory rate <30/min, and no shock or organ dysfunction, c) Severe: FiO_2_ 32-40%; PaO_2_ / FiO_2_ ratio ≥ 200, respiratory rate ≥ 30/min, without shock or organ dysfunction, and d) Critical: FiO_2_> 40%; PaO_2_ / FiO_2_ ratio <200, septic shock, organ dysfunction, chest x-ray with bilateral infiltrates and / or requirement for invasive mechanical ventilation (IMV). Patients in severe or critical conditions were transferred to the Regional Hospital. The guideline considered the use of hydroxychloroquine (HQ; day 1: 400mg every 12hrs, day 2-5: 200mg every 12 hrs) in all admitted patients as well as those > 60 years and / or with chronic diseases. All admitted patients received a beta-lactam antibiotic and pharmacological prophylaxis for thromboembolic disease. Besides, critical patients with a D-dimer (DD) value > 6 times the normal value were prescribed with an intermediate heparin dose (unfractionated formulation 5,000 UI every 8 hours or enoxaparin 1 mg/kg/day). The use of glucocorticoids and convalescent plasma were indicated according to the criteria of physicians in charge.

### Clinical characterization and outcomes

Demo-epidemiological variables, pregnancy status and comorbidities as well as clinical features were retrieved from medical records. The CALL score was calculated as described (21) as well SOFA and APACHE II in those admitted to the ICU. Pharmacological therapies, complications, clinical outcomes, length of stay and in-hospital mortality were extracted and calculated from medical records.

### Statistical analysis and ethical issues

Data corresponding to quantitative variables is presented as medians with interquartile range (IQR) and categorical variables as percentages. Analysis of potential factors associated with ICU admission was performed with non-parametric tests such as the Fisher or the Mann-Whitney test. Some graphs were fitted with the least squares method to facilitate trends reading. This work was approved by the Institutional Review Board.

## Results

### Cases identified and epidemiological features

Twenty-one patients were admitted to our hospital between epidemiological weeks 13 and 20, a group that compose this preliminary report. During this timeframe, hospital admissions at our center (all in severe or critical condition) represented 8.8% of the total reported cases in the region (21 out of 237). Figure 1 shows the impact of the first wave on admissions at our reference center. Admissions (to all units or ICU) decreased secondary to mitigation measures.

**Figure 1.**
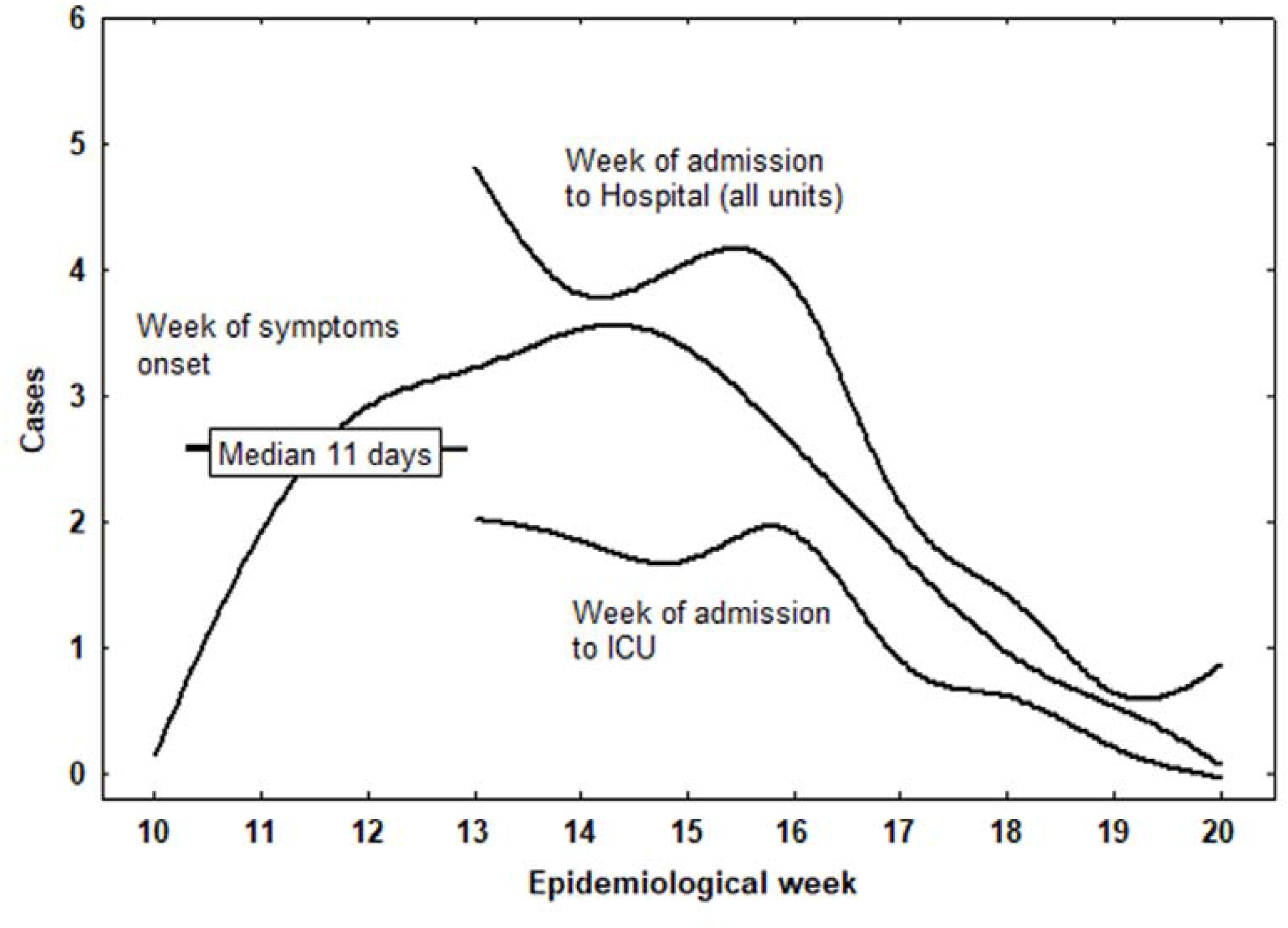
Distribution of cases with Covid-19 admitted to the hospital according to the epidemiological week from symptoms onset to the actual week of hospitalization. More than a week of delay between symptoms onset and admission is observed. Curves were fitted by the least square method

### Clinical features

Median age was 61 years (range 26-85 years, IQR 51-68 years) and more than half were ≥ 60 years old (Table 1). Female patients predominated and contact with a known Covid-19 case was frequent. A healthcare worker integrates this report whose contagion occurred in a primary care center. Hypertension, obesity, and type 2 diabetes mellitus were prevalent (Table 1). Two patients were pregnant and 19% had no comorbid history. The use of active drugs on the renin-angiotensin-aldosterone system approached 40% of the series (Table 1). Predominant symptoms in order of frequency were cough, dyspnea, and fever followed by myalgia, odynophagia, and headache. Anosmia and dysgeusia were reported in about a quarter of patients. Diarrhea and rhinorrhea were infrequent but observed in about a fifth of cases (Figure 2). Arterial hypotension on admission was infrequent (9.5%). The median symptomatic period before admission was 11 days (IQR 5-13 days, Figure 1). About half of our patients were admitted with tachypnea and an abnormal PaO_2_ / FiO_2_, but low O_2_ saturation on pulse oximetry or hypoxemia in arterial gases was less frequent (Table 2). We did not observe patients with leukocytosis or neutrophilia. In contrast, the fraction of patients with lymphopenia (42.9%) and to a lesser degree, thrombocytopenia (23.8%) was relevant. The series also highlights the high frequency of patients with elevated LDH and ferritin plasmatic values (> 70%, Table 2). Coagulation parameters (International Normalized Ratio and activated partial thromboplastin time) did not show major alterations, but DD values were elevated in about half of patients.

**Table 1.**
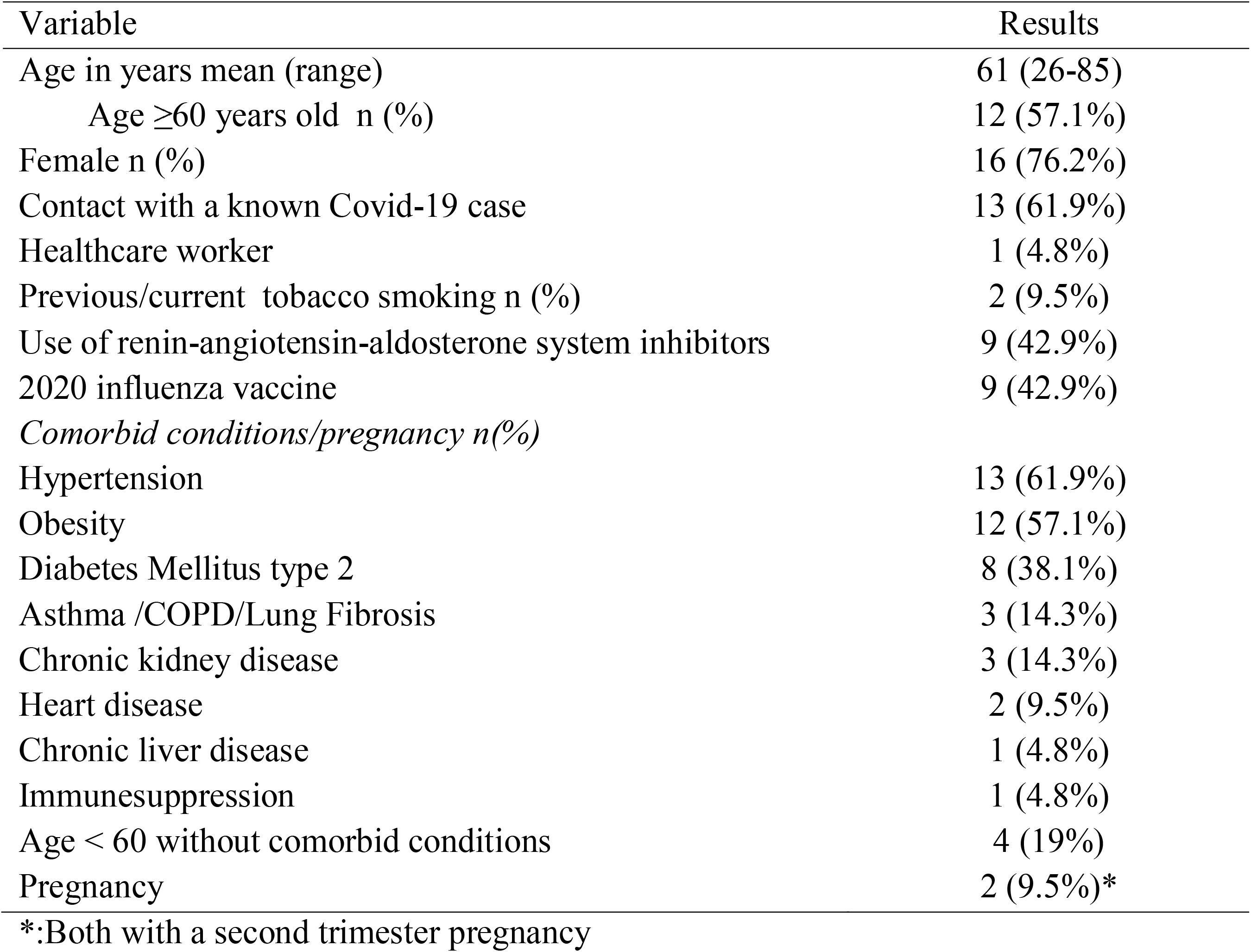
Features of 21 patients admitted with Covid 19 at a Regional Reference Center in Valdivia, southern Chile during the first pandemic wave 2020.

| Variable | Results |
| --- | --- |
| Age in years mean (range) | 61 (26-85) |
| Age $\geq 60$ years old n (%) | 12 (57.1%) |
| Female n (%) | 16 (76.2%) |
| Contact with a known Covid-19 case | 13 (61.9%) |
| Healthcare worker | 1 (4.8%) |
| Previous/current tobacco smoking n (%) | 2 (9.5%) |
| Use of renin-angiotensin-aldosterone system inhibitors | 9 (42.9%) |
| 2020 influenza vaccine | 9 (42.9%) |
| *Comorbid conditions/pregnancy n(%)* |  |
| Hypertension | 13 (61.9%) |
| Obesity | 12 (57.1%) |
| Diabetes Mellitus type 2 | 8 (38.1%) |
| Asthma /COPD/Lung Fibrosis | 3 (14.3%) |
| Chronic kidney disease | 3 (14.3%) |
| Heart disease | 2 (9.5%) |
| Chronic liver disease | 1 (4.8%) |
| Immunesuppression | 1 (4.8%) |
| Age < 60 without comorbid conditions | 4 (19%) |
| Pregnancy | 2 (9.5%)* |

\*:Both with a second trimester pregnancy

**Table 2.** Physical and laboratory parameters at admission among 21 patients admitted with Covid 19 at a Regional Reference Center in Valdivia, southern Chile during the first pandemic wave.

| Parameter | Results |
| --- | --- |
| Temperature °C median (IQR) | 37.1 (36.6-37.8) |
| No fever | 4 (19%) |
| Respiratory rate /min median (IQR) | 20 (25-34) |
| Tachypnea ($\geq 30$/min) n (%) | 10 (47.6%) |
| Heart rate /min mean (IQR) | 102 (81-115) |
| O<sub>2</sub> pulse digital oxymetry % median (IQR) | 94% (91-96) |
| O<sub>2</sub> Saturation <90% n (%) | 4 (19%) |
| PaO<sub>2</sub> at admission mmHg median (IQR) | 76.4 (69.9-90.4) |
| PaO<sub>2</sub> <60 mmHg | 1 (4.8%) |
| PaFiO<sub>2</sub> median (IQR) | 295 (205-375) |
| PaFiO<sub>2</sub> <300 n (%) | 12 (57.1%) |
| Blood leucocyte count/μL median (IQR) | 6730 (6310-8690) |
| Leucocytosis >15000 /μL | 0 (0%) |
| Blood polymorphonuclear count/ μL median (IQR) | 5400 (4700-6800) |
| Neutrophilia >12000/μL | 0 (0%) |
| Lymphocyte count /μL median (IQR) | 1200 (900-1400) |
| Lymphocyte count $\leq 1000$/ μL n (%) | 9 (42.9%) |
| Platelet count /μL median (IQR) | 209000 (183000-306000) |
| Thrombocytopenia < 100000 /μL median n (%) | 5 (23.8%) |
| LDH in plasma U/L median (IQR)* | 305 (238-467) |
| $\leq 250$ U/L (normal range) | 6 (28.6%) |
| 250-500 U/L | 12 (57.1%) |
| > 500 /UL | 1 (4.8%) |
| Blood urea nitrogen mg/dL median (IQR) | 16 (10-21) |
| C reactive protein mg/L median (IQR) | 9.6 (7-14.4) |
| International normalized Ratio median (IQR) | 1.12 (1.06-1.16) |
| Activated partial thromboplastin time median (IQR) sec | 30.4 (29-40.8) |
| D dimer μg/mL median (IQR) | 0.63 (0.33-1.58) |
| D dimer over normal value (> 0.5 μg/mL) | 10 (47.6%) |
| Plasmatic ferritin ng/mL | 552.7 (370.4-1168) |
| Over normal value (> 150 ng/mL) | 18 (85.7%) |
| > 800 ng/mL | 6 (28.6%) |

\*: of patients with available results

**Figure 2.**
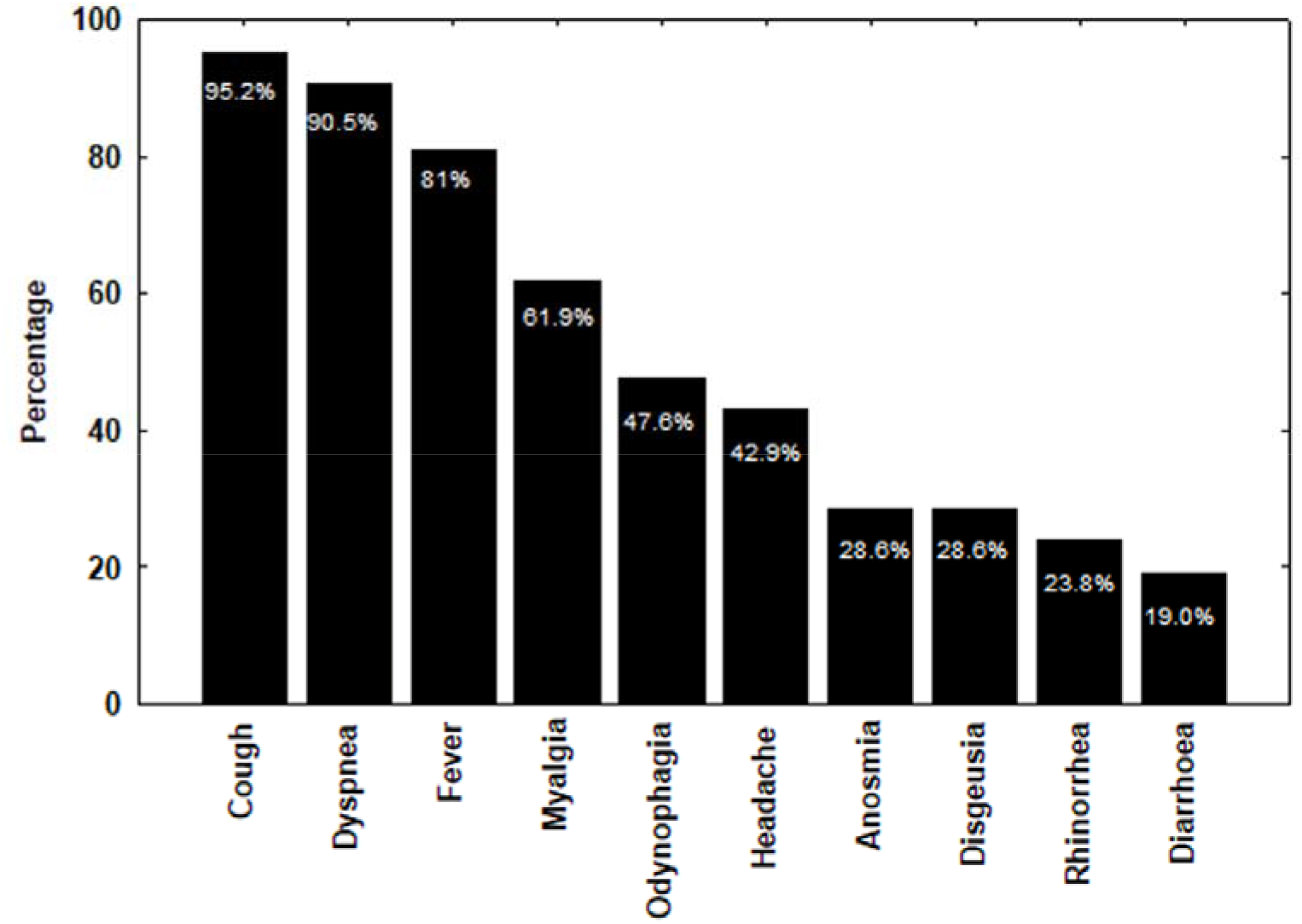
Distribution of symptoms among 21 patients admitted to a Regional Reference Center with Covid 19 in Valdivia, Southern Chile.

### Management, complications and outcome

Approximately 40% (9/21) of patients were transferred to ICU (Table 3), all requiring IMV with median SOFA and APACHE II scores of 4 points (range 2-9) and 12 points (range 3-20), respectively. Half of the patients had a CALL score in the highest severity range (10-13 points, Table 3). The use of vasoactive drugs was required in 8 of the 9 patients with IMV. All the patients received by protocol, HQ and about half also azithromycin. Furthermore, 5 of the 21 patients received steroid therapy (23.8%, Table 3). Corticosteroids were indicated in patients with refractory ARDS (methyl prednisolone), refractory shock (hydrocortisone), obstructive bronchitis (hydrocortisone), thrombocytopenic purpura (dexamentasone) and to achieve fetal maturation (betamethasone) in a pregnant woman. In the latter case, she had a 28 weeks pregnancy and underwent an urgent cesarean section, receiving the only convalescent plasma therapy in the series shortly before delivery. She had a premature newborn weighing 1540 grams, with hypotonia, intraventricular hemorrhage, seizures, neonatal jaundice and later bacteremia, requiring IVM connection and application of pulmonary surfactant. Perinatal transmission of SARS-CoV-2 was ruled out. Until the time of publication, he was still hospitalized in the neonatal ICU.

**Table 3.** CALL score, management, complications and outcomes among 21 patients admitted with Covid 19 at a Regional Reference Center in Valdivia, southern Chile during the first pandemic wave.

| Variable | Results |
| --- | --- |
| CALL score n (%) |  |
| 4-6 points | 3 (14.3%) |
| 7-9 points | 7 (33.3%) |
| 10-13 points | 11 (52.4%) |
| Place of hospitalization n (%) |  |
| General ward | 12 (57.1%) |
| Critical Care Unit | 9 (42.9%) |
| Ventilatory support n (%) |  |
| Invasive mechanical ventilation | 9 (42.9%) |
| Prone position | 6 (28.6%) |
| Non-invasive mechanical ventilation only | 0 (0%) |
| High-flow nasal cannula | 7 (33.3%) |
| Vaso-active drugs | 8 (38.1%) |
| Drug Treatment n (%) |  |
| Hydroxychloroquine | 21 (100%) |
| Macrolides | 10 (47.6%) |
| High-intermediate doses of low-molecular weight heparin | 6 (33.3%) |
| Corticosteroids | 5 (23.8%) |
| Convalescent plasma | 1 (4.8%) |
| Cesarean section | 1 (4.8%) |
| Complications n (%) |  |
| ARDS | 9 (42.9%) |
| Long QTc interval* | 1 (4.8%) |
| Acute Kidney Injury** | 6 (28.6%) |
| Hospital-acquired infections | 7 (33.3%) |
| Hospital length of stay |  |
| Median (IQR) in days | 18 (10-28) |
| ≥ 14 days n (%) | 12 (57.1%) |
| Deceased n (%) | 2 (9.5%) |

\*. No arrhythmia associated; \*\*one case with dialysis requirement.

Among observed complications, Acute Respiratory Distress Syndrome (ARDS) was recorded in 9 patients (42.9%), nosocomial infections in 33.3%, and Acute Kidney Injury in 28.6% (with dialysis requirement in one of them). An isolated prolonged QTc interval event was registered but without arrhythmia (Table 3). Health care-associated infections included 3 events of bloodstream infections, 2 of ventilator-associated pneumonia, 1 of urinary tract infection, and one of fever without an identified source. Median hospital stay was 18 days with more than half remaining hospitalized ≥ 2 weeks. Two of our patients died (in-hospital mortality 9.5%) at 35 and 38 days after admission due to central venous catheter-associated bloodstream infection and refractory ARDS, respectively.

### Serological and molecular studies

All patients underwent a rapid serologic test to detect IgM / IgG antibodies. After the first round (taken at different weeks of symptoms onset), most showed IgM+/IgG + (81%) or isolated IgM+ (9.5%) antibodies (Table 4). Negative and invalid results were also observed. A second round of examinations carried out in the 4 patients with negative, invalid or isolated positive IgM results, demonstrated seroconversion to IgM+/IgG+ in all of them. Rapid test sensitivity during the first round increased according to the elapsed time but was low during the first 3 weeks (19% to 61.9%, Table 4, Figure 3).

**Table 4.**
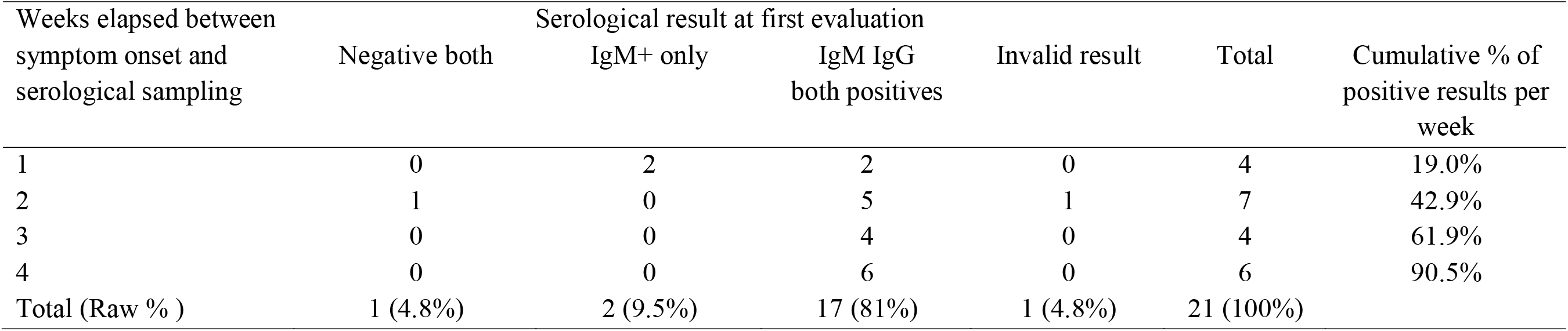
Distribution of results after the first serological evaluation of IgM and IgG anti-SARS-CoV2 antibodies.

**Figure 3.**
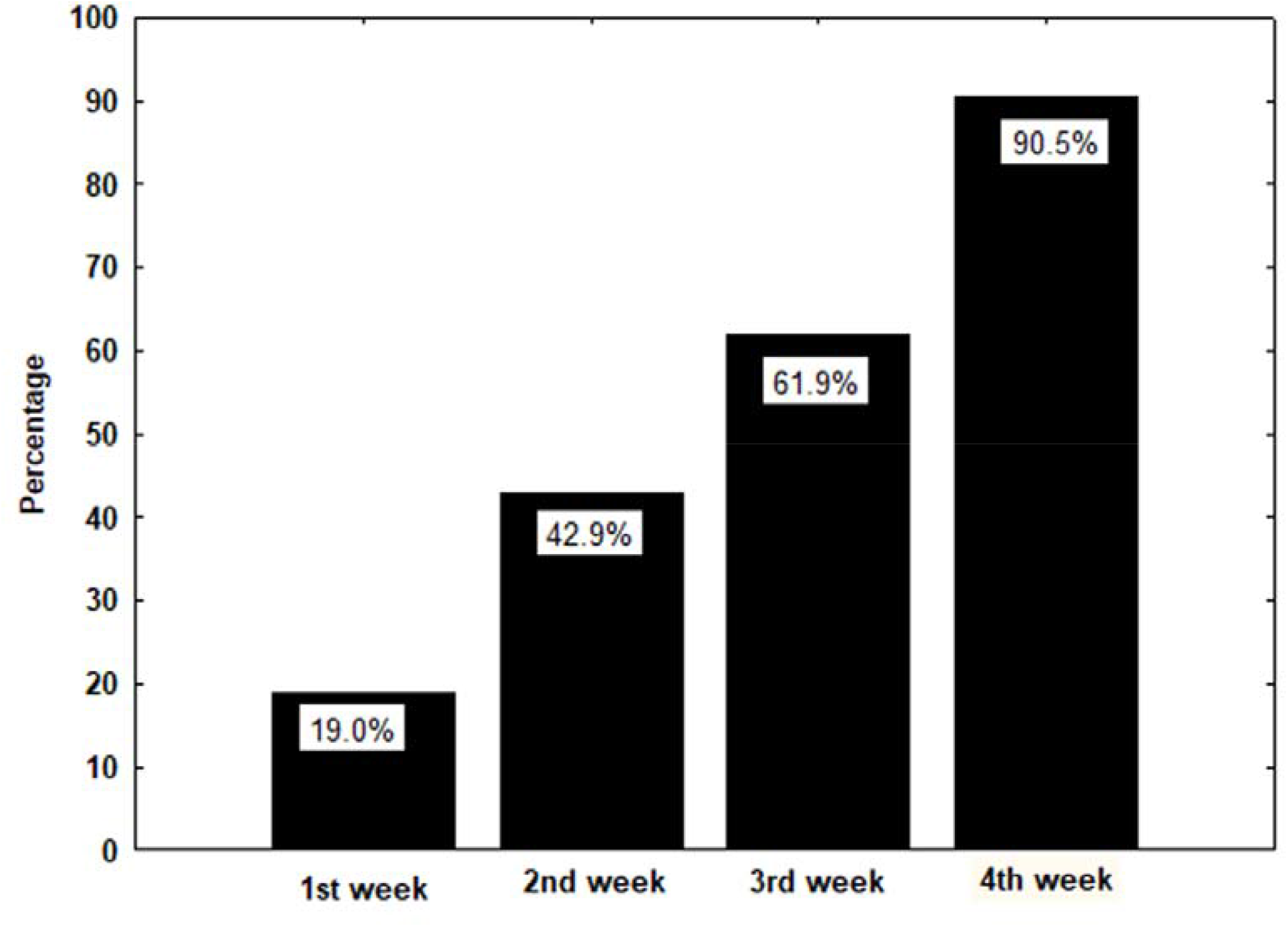
Accumulated sensitivity per week of symptoms onset after the first serological evaluation of IgM and IgG anti-SARS-CoV2 antibodies using a Rapid commercial test. Either a positive IgM or a combined IgM/IgG positive result was considered for analysis.

### Comparison of critical versus non-critical group

An exploratory analysis was performed to identify possible factors associated with a transfer to ICU. A critical evolution was not associated with increased age, gender, comorbidities, pregnancy, or a higher CALL score (data not shown). However, 2 factors appeared linked to ICU admission: tachypnea and elevated plasma LDH values. Seven out of 9 patients with a critical condition had tachypnea vs. 3 out of 12 without a critical condition (OR 10.5; IC95 1.36-81; p <0.05). Plasmatic LDH values in the critical group had a median of 467 U/L (IQR 427-493) versus 267.5 U/L (IQR 238-305) in the non-critical group (p <0.05). Due to the small size of the groups, a multivariate analysis was not attempted.

## Discussion

After attacking China, Europe, and the United States, the epicenter of the Covid-19 pandemics is centered currently in Latin America. However, there is little information on the clinical features, management, and outcome for this infection in our region. This work contributes to the knowledge of this infection in the Latin American context and communicates its preliminary results. Patients were managed using a protocol built according to the best evidence available at the time of its preparation (22-25) and although the pharmacological therapies initially contemplated are currently questioned or disregarded (26-30), the existence of a guideline made it possible to work in a uniform manner in a whole region of southern Chile.

As in other experiences, we were able to confirm that hospital admissions occur several days after the onset of symptoms, that less than 10% of all cases require hospitalization and that hospital stay is prolonged (10,17). Patient characteristics reflect the weight of comorbidities and advanced age, involvement of healthcare personnel or pregnant women, and known systemic and respiratory symptoms in addition to changes in taste or smell and occasionally gastrointestinal manifestations. In our series, ICU admission for critical conditions was very high (about 50% of admissions) because our center concentrates severe and critical patients of the region. All patients received HQ per protocol, a drug whose efficacy and safety has recently been questioned (26, 27, 31). In our series, only one case of QTc prolongation was observed and none presented arrhythmias.

Steroid therapy was applied on a case-by-case basis for different reasons and not by protocol, since there is no consensus regarding its use. Although the available evidence has not been consistent (32, 33), some experiences have suggested that administration could increase survival (34-36). Its use in turn, must be balanced with its known association with infectious complications, myopathy, delirium, hyperglycemia, gastrointestinal bleeding or hydro-electrolyte disorders (37). ARDS secondary to other viral etiologies have been associated with a delayed viral clearence (38) or even increased mortality (33, 39). Currently, some prospective projects are underway that could solve these questions.

There were 2 patients carrying a second trimester pregnancy who presented with pneumonia and respiratory failure due to SARS-CoV-2, one of them with severe illness that responded to conventional O_2_ therapy without fetal or maternal complications. The second evolved in a critical condition, requiring IMV, and prompt fetal lung maturation with corticosteroids. Fortunately her evolution was favorable after an urgent cesarean section and convalescent plasma therapy. The impact of this infection in pregnancy is beginning to be delineated with no intrauterine transmission events or maternal mortality so far reported and with most of the deliveries managed by Cesarean sections (40). Fetal consequences may include intrauterine growth retardation, cyanosis, gastrointestinal bleeding, and neonatal mortality (40).

Convalescent plasma is an experimental potentially beneficial therapy (41, 42) that has shown an adequate safety profile (43). Recently, a randomized clinical trial reported a trend towards a higher proportion of patients with clinical improvement (28 days) and lower mortality in the plasma-receiving group (44).

Laboratory tests showed an inflammatory state (elevated C-reactive protein, ferritin and plasmatic LDH levels) and procoagulant status (increased DD). The relevance of tachypnea and elevated plasma LDH in the identification of critically ill patients was confirmed. These results are consistent with other studies where increased LDH values had a high sensitivity (100%) and specificity (86.7%) to predict severe disease (45). In our report, the low number of cases prevented confirming other variables associated with severity or adverse prognosis such as advanced age, high fever, chronic diseases (especially chronic cardiovascular or pulmonary disease, diabetes and high blood pressure), coagulopathy, increased D-dimer, troponin or ferritin levels, neutrophilia, lymphopenia, and/or leukocytosis (34, 46-49).

Serology by a rapid antibody-based test was performed in all patients with 19% and 42.9% sensitivity in the first and second week, respectively, consistent with other reports (50, 51) that reinforce its limited utility for early diagnosis. However, it can be helpful when a high suspicion exists despite serial negative RT-PCT and compatible symptoms and chest-CT images as happened in one of our patients.

Complications observed underline the high frequency of ARDS and healthcare associated infections which in turn explained death of our patients (52). Autopsy studies have revealed the presence of diffuse alveolar damage, in exudative and proliferative phases, including alterations such as capillary congestion, pneumocyte necrosis, hyaline membrane, interstitial edema, atypical and hyperplastic pneumocytes, and microthrombi in arterioles (53, 54). The latter support using anticoagulants in patients with high DD (55) which in observational studies has shown to reduce mortality (56, 57). Thus all our patients received heparin prophylaxis and in one third we use high-intermediate doses due to high DD values.

## Conclusions

In our regional Center, Covid-19 was associated to known risk factors, had a prolonged stay and in-hospital mortality. Near 8% of regional cases evolved with a severe or critical illness and required to be transferred to a designated reference hospital. Tachypnea ≥30/min and high plasmatic LDH levels were predictive of transfer to ICU, mainly by ARDS. Rapid antibody-based tests were not useful for early diagnosis. Besides ARDS, hospital-acquired infections and acute kidney injury were frequent complications. As specific therapeutic alternatives have no proven effectiveness, Covid-19 management rest mainly on supportive care.

## Data Availability

Data is available if needed

